# Relationships among Commercial Biases and Author Conflicts of Interest in Biomedical Publishing

**DOI:** 10.1101/2020.01.24.20018705

**Authors:** S. Scott Graham, Zoltan Majdik, Dave Clark, Molly M. Kessler, Tristin Brynn Hooker

## Abstract

Recently, concerns have been raised over the potential impacts of commercial biases on editorial practices in biomedical publishing. Specifically, it has been suggested that commercial biases may make editors more open to publishing articles with author conflicts of interest (aCOI). Using a data set of 128,781 articles published in 159 journals, we evaluated the relationships among commercial publishing biases and reported author conflicts of interest. The 159 journals were grouped according to commercial biases (reprint services, advertising revenue, and ownership by a large commercial publishing firm). 30.6% (39,440) of articles were published in journals showing no evidence of commercial publishing biases. 33.9% (43,630) were published in journals accepting advertising and reprint fees; 31.7% (40,887) in journals owned by large publishing firms; 1.2% (1,589) in journals accepting reprint fees only; and 2.5 % (3,235) in journals accepting only advertising fees. Journals with commercial biases were more likely to publish articles with aCOI (9.2% (92/1000) vs. 6.4% (64/1000), p = 0.024). In the multivariate analysis, only a journal’s acceptance of reprint fees served as a significant predictor (OR = 2.81 at 95% CI, 1.5 to 8.6). Shared control estimation was used to evaluate the relationships between commercial publishing biases and aCOI frequency in total and by type. BCa-corrected mean difference effect sizes ranged from -1.0 to 6.1, and confirm findings indicating that accepting reprint fees may constitute the most significant commercial bias. The findings indicate that concerns over the influence of industry advertising in medical journals may be overstated, and that accepting fees for reprints may constitute the largest risk of bias for editorial decision-making.

## Introduction

For some time now, there has been growing concern about the extent to which financial relationships with industry bias the results of biomedical research. Studies of industry funding and author conflicts of interest (aCOI) in the biomedical sciences have found that these financial relationships can bias choices in experimental design^1-3^ as well as clinical decision-making during trial execution.^4-6^ In particular, the most recent studies and meta-analyses confirm that these financial relationships and associated practices result in a substantial increase in the likelihood that clinical trial results will be favourable to industry.^7-9^ Recent research in these areas also points toward an ever-widening array of potentially biasing practices, including ghost authorship^10-11^ and so-called “marketing trials”—i.e., clinical trials that were designed primarily to influence medical decision-making in favour of product use.^12-13^ Despite movements toward greater transparency in disclosing aCOI in medical journals, including the International Committee for Medical Journals Editors (ICJME) recommendations for reporting aCOI,^14^ inconsistencies still remain in reporting financial and non-financial COI for authors, researchers, and editors. This issue is particularly acute regarding the relative inconsistency and opacity of editorial COI disclosures, a concern that often persists even when author and researcher disclosures become more transparent.^15-18^

In addition to worries over personal COI that may be held by journal editors, there are also growing apprehensions over the potential effects of commercial publishing biases on biomedical research. Specifically, it has been suggested that journal-level financial relationships such as the acceptance of industry advertising revenue, reprint fees, and additional industry printing contracts held by journal parent companies may impact editorial decision-making, creating an environment more favourable to industry-sponsored research.^19-21^ Editor COI and commercial publishing biases may be of particular concern given recent fears that non-peer-reviewed publications with aCOI are having significant impacts on biomedical research and clinical practice.^25^ Certainly, available anecdotal evidence does suggest there may be cause for concern.^22-23^ Two of the most notable cases involve the punitive withdrawal of $1.5 million in advertising revenue from the *Annals of Internal Medicine* following the publication of an article critiquing multiple industry-funded trials in 1992^20^ and Merck’s dispersal of $836,000 to the *New England Journal of Medicine* for reprints of the VIGOR study as a part of the Vioxx marketing campaign.^24^ Furthermore, there are some data available indicating that commercial publishing biases may lead to editors being less diligent in the execution of journal aCOI policies for article authors.^19-20^ As greater attention is paid to the potential adverse consequences of aCOI and industry funding on medical research, it is critical that ongoing discussions regarding commercial publishing biases occur in an evidence-rich environment.

Accordingly, this study evaluates the relationships between commercial publishing biases and industry favourability (as measured by aCOI likelihood and frequency) in 159 biomedical journals. Since aCOI have consistently and reliability been shown to result in research results favourable to industry, they serve as an effective surrogate endpoint for measuring industry favourability broadly. In what follows, we describe our development of an AI framework for identifying and classifying aCOI. We then compare aCOI likelihood and frequency in journal samples stratified by identified commercial biases. As the results described below detail, the presence of some commercial biases does appear to create an environment more favourable to scholarship with aCOI, increasing the likelihood that published articles will have aCOI as well as the number of aCOI per article. In particular, the acceptance of reprint orders appears to the most influential of evaluated commercial biases.

## Methods

In order to enhance the available evidence base regarding the potential influence of commercial publishing biases on editorial decision-making, we collected 128,781 biomedical journal articles indexed with Medline. In 2016 Medline began collecting conflicts of interest information from participating journals. We extracted data for analysis in January 2019, and at that time, approximately 30 million articles were indexed in the database. The population of 128,781 articles was identified first by extracting all MEDLINE-indexed articles (2016-2018) with aCOI disclosure statements (N=274,246). These articles were published in a total of 1497 journals. Our final sample of 128,781 articles in 159 journals was derived by excluding all articles where the publishing journal was present fewer than 25 times in the full dataset. We evaluated the presence and rate of aCOI across all articles in the dataset using a custom-built automated parser and compared aCOI likelihood and quantity to measures of commercial biases. In what follows, we describe our approaches to 1) aCOI identification and classification, 2) evaluating the reliability of the aCOI parser, and 3) identifying the presence or absence of commercial publishing biases in each journal.

### Author COI Identification & Classification

In order to identify and classify each of the reported aCOIs in these disclosure statements, we developed a metadata assisted, machine-learning enhanced, natural language processing (NLP) tool. In short, the parser uses a trained language model to tag sponsors (e.g., pharmaceutical companies). The parser then uses Medline author metadata to identify named authors in the disclosure statements, matches authors to sponsors, and finally identifies the type of conflict disclosures. Each of these parser stages are described in more detail below.

#### Sponsor Identification

An NLP method called Named Entity Recognition (NER) can reliably use grammatical and/or statistical techniques to extract and classify entities like persons, locations, dates, or organizations from unstructured text. For example, a sentence such as “Walter Sandulli and Jessica Goldenberg are employees of Akrimax,” when parsed, would produce three “named entities”: Walter Sandulli, PERSON; Jessica Goldenberg, PERSON; and Akrimax, ORG. NER approaches can work accurately on unknown texts, and can achieve near-human levels of precision when trained using a machine learning approach. In the case of disclosure statements, the lack of consistent styling in the writing and editing of COI statements means that organization names are presented very differently, sometimes within the same COI statement (e.g., GlaxoSmithKline vs. Glaxo vs. GSK). COI statements are similarly inconsistent in presenting author names; often they use initials, but sometimes last names or other abbreviations will be present. Using this custom-trained language model based on the small sample of human-corrected COI statements (n=100), we were able to decrease the sponsor identification error rate by 68%.

#### Author Identification

Our approach used MEDLINE data on author names to further increase recognition accuracy for both author names and organizations. In light of the author naming conventions described above, as well as the fact that organizations in the biomedical field often have names that, to a computer, resemble human names (e.g., the “Smith Kline” of GlaxoSmithKline), automated NER parsing will frequently mischaracterize organizations as names, and vice versa. To counteract this issue, the parser uses author metadata to generate an author-name permutation table with 13 name permutations that correspond to author naming conventions from various journal style guides for disclosure statements. “Jane Alicia Doe,” for example, would be rendered as “J.A.D.,” “J. Doe,” “J Doe,” and ten other permutations of first, middle, and last name and/or initials. Using this metadata-generated list of author permutations instead of relying on the NER to tag both authors and organizations allowed us to not only have a high degree of precision in identifying authors in COI statements, but also to cross-check them against entities tagged as organizations and remove them if they were in the author list.

#### aCOI Classification

The aCOI classification dictionary is based loosely on the International Committee of Medical Journal Editors (ICMJE) standardized conflicts of interest disclosure form. Our COI dictionary schema organizes these categories (as well as employment in industry) into a three-level schema based on potential benefit from a product’s success. Specifically, low-level aCOI included personal fees, travel, board memberships, and non-financial support. Mid-level aCOI included grants and research support. Finally, high-level aCOI included stock ownership and employment in industry. The parser assumes a standard syntax that almost all COI disclosure statements follow, where a name (or names) are followed by an aCOI disclosure type (like “is employed by”), which is followed by the aCOI source. The parser extracts aCOI value(s) from each disclosure statement by stitching the three elements described above—NER, author permutations, aCOI classifications—together through a regular expression. This process is repeated for each tagged sponsor in a disclosure statement. Outputs are collated and assigned a numerical weight based on the aCOI classification dictionary. Table 1 provides an example of a fully parsed disclosure statement.

**Table 1:** Fully-parsed disclosure statement for “Defining Priorities for Future Research: Results of the UK Kidney Transplant Priority Setting Partnership” (PMID: 27776143).

| Author | Relationship Type | Sponsor | Conflict Weight |
| --- | --- | --- | --- |
| Simon Ball | grant | Oxford | 2 |
| Simon Knight | fees | OrganOx UK | 1 |
| Lorna Marson | fees | Novartis | 1 |
| Lorna Marson | fees | Astellas | 1 |
| Fiona Loud | fees | Merck | 1 |
| Graham Lipkin | fees | Raptor Pharmaceuticals | 1 |
| Graham Lipkin | fees | Alexion Pharma | 1 |

#### Parser Reliability

In order to evaluate the reliability of the aCOI parser, a random sample of 1000 disclosure statements was submitted to human evaluation. While the dataset includes 128,781 disclosure statements, the results of our analysis indicate that approximately 94% of these are some version of “The authors report no conflicts of interest.” A truly representative sample of 1000 disclosure statements would thus only provide 60 statements for the human or parser to evaluate. Therefore, our sampling protocol excluded disclosure statements of fewer than 50 characters (i.e., those more likely to be some variation of “The authors report no conflicts of interest,” which is 44 characters long. The end result of this approach is that we oversampled disclosure statements where aCOIs were more likely to be present.

In order to compare the human-coded and machine-coded samples, we assessed reliability using the two-way average measure Intra-Class Correlation Coefficient (ICC). The average ICC for low-level conflicts was 0.722, with a 95% confidence interval from 0.69 to 0.751 (F[998,903[= 6.27, p < .01). The average ICC for medium weight conflicts was 0.773, with a 95% confidence level from 0.747 to 0.797 (F[998,985] = 7.84, p < .01). And, finally, the average ICC for high-level conflicts was 0.618, with a 95% confidence level from 0.578 to 0.656 (F[998,923] = 4.28, p < .001).

Recommendations for appropriate ICC thresholds vary somewhat across disciplines and contexts. The threshold of “low” agreement can be from below ICC = 0.40^26^ to ICC = 0.50.27 Fair to moderate agreement thresholds vary the most with recommend ranges from ICC= 0.40 to ICC = 0.75.^28^ Most ICC schemata accept ICC > 0.60 as fair to good and ICC > 0.75 as good to excellent. Since identifying the absence of conflicts is an easier computational task than conflict classification, our approach here invariably resulted in lower ICC scores than would be expected in a truly representative sample. However, the benefit of this approach is that it ensured the parser evaluation would involve a much wider variety of conflict types. Nevertheless, parser reliability scores generally fell within ranges that would be classified as moderate to good.

### Commercial Biases Identification

Potential sources of commercial bias were identified based on the extant literature. Research and opinion pieces published in biomedical journals regularly identifies the acceptance of advertising revenue, the acceptance of reprint contracts, and the parent company’s acceptance of industry publishing contracts (e.g. supplements) as potential sources of editorial bias.^20,34^ Therefore, we reviewed journal websites for solicitations of adverting revenue and reprint fees. Additionally, for each journal, the parent company of the journal was identified. This information is typically available in a website header or footer and/or on the “About” page. If the parent company was primarily a publishing firm (e.g. Elsevier, Taylor and Francis, Wiley), the journal was assigned to the large publishing firm category. Every journal in the dataset that was owned by a large publishing firm accepted adverting and reprint fees. Thus we were able to assign each journal to one of the following categories: 1) control group (accepts no advertising, reprint fees, not owned by commercial publishing group); 2) accepts advertising revenue, but not reprint fees, 3) accepts reprint fees, but not adverting revenue, 3) accepts both advertising and reprint fees, but is not owned by a large commercial publisher, and 4) owned by a commercial publishing firm.

## Results

We evaluated aCOI rates for 128,781 articles published in 159 journals indexed by MEDLINE. Each journal in the dataset included at least 25 articles, with *PLoS One* having the most at 22,252 articles. By group, the dataset included 43,630 articles in journals accepting advertising and reprint fees, but not belonging to a commercial publishing firm; 40,887 articles in journals owned by large publishing firms; 1,589 in journals accepting reprint fees but not advertising fees; 3,235 in journals accepting only advertising fees; and 39,440 articles in the control group. Table 2 details these numbers alongside aCOI rates.

**Table 2:** Number of articles and aCOI rates by journal commercial bias category

| Group | n | Low aCOI | Mid aCOI | High aCOI | Total aCOI |
| --- | --- | --- | --- | --- | --- |
| Control | 39440 | 6912 | 4960 | 6451 | 18326 |
| Reprints | 1589 | 982 | 750 | 242 | 1974 |
| AdRev | 3235 | 600 | 266 | 1975 | 1147 |
| AdRev + Reprints | 43630 | 34425 | 24741 | 6180 | 65346 |
| CommPub | 40887 | 20834 | 12924 | 6806 | 40564 |

**Table 3:**
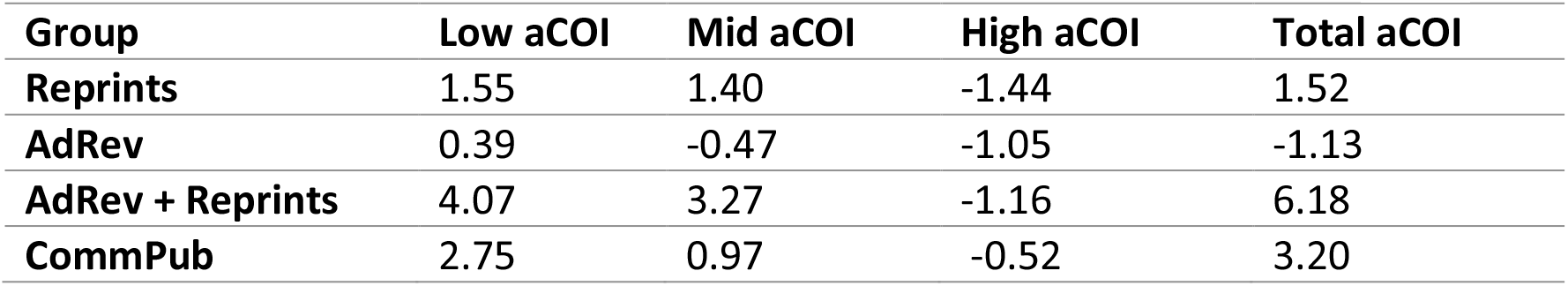
Effect size expressed as BCa-corrected (5000 iterations) mean difference compared to the control

| Group | Low aCOI | Mid aCOI | High aCOI | Total aCOI |
| --- | --- | --- | --- | --- |
| Reprints | 1.55 | 1.40 | -1.44 | 1.52 |
| AdRev | 0.39 | -0.47 | -1.05 | -1.13 |
| AdRev + Reprints | 4.07 | 3.27 | -1.16 | 6.18 |
| CommPub | 2.75 | 0.97 | -0.52 | 3.20 |

### aCOI Frequency Analysis

An initial test for equality of proportions (using Yates’ continuity correction) was conducted to assess if articles with aCOI were more likely to be published journals with commercial biases. In order to ensure adequate statistical power (β=.9) for the test, a random sample of 1,000 articles was selected from each set of journals (with vs. without commercial biases). Journals with commercial biases published articles with aCOI at a rate of 9.2% whereas those without commercial biases published articles with aCOI at a rate of 6.4%. This difference is significant [χ^2^ (1, N=2000) = 5.07, p= 0.024].

In order to evaluate whether some commercial biases were more predictive of these frequency differences, we identified the rate at which articles with aCOI were published in each journal in the data set. These data were fitted to a quasi-binomial multiple regression model with a logit link. Overall, the model was significant at F(23.33, 2496)=3.51, p=0.017. Neither advertising revenue nor ownership by a large commercial publishing firm were significant predictors of aCOI likelihood. However, a journal’s acceptance of reprints predicts that the likelihood that a published article will have aCOI increases by a factor of 2.81(95% CI, 1.5 to 8.6, p=0.0416).

### aCOI Rates Analysis

Shared-control estimation plots were used to compare aCOI rates across journal categories. Shared-control estimation plots are part of the estimation statistics framework recently promulgated as a robust alternative to null-hypothesis significance testing.^29^ Estimation plots focus analytic attention on population parameters, mean differences, and effect sizes over p-values. The approach here uses Efron’s technique for bias-corrected accelerated bootstrap (BCa) estimation to account for skewed populations.^30^ Using a stratified sample of articles in each group, we ran 5,000 BCa iterations at the 95% confidence level in order to derive the effect estimates reported below.

The results of the estimation plots (Figure 1) indicate that commercial biases have a modest effect on the number of aCOI per article. Of course, these effects are not equal across categories or measures. In terms of total aCOI, the effect size is so modest as to be negligible for journals accepting either reprints or ad revenue. However, for journals accepting both reprints and ad revenue the effect size would functionally double the aCOI rate for the average article, whereas ownership by a large commercial publishing firm would increase the aCOI rate of the average article by approximately 60%. However, when aCOI are separated by weight, a more complex picture emerges. Among the aCOI-level specific plots, the largest effect sizes appear on the plot for low-level aCOIs. Interestingly, all the effects are negative on the plot for high-level aCOIs. However, the findings for high-level aCOI should be interpreted cautiously given the more moderate inter-rater agreement rates.^31^

**Figure 1:**
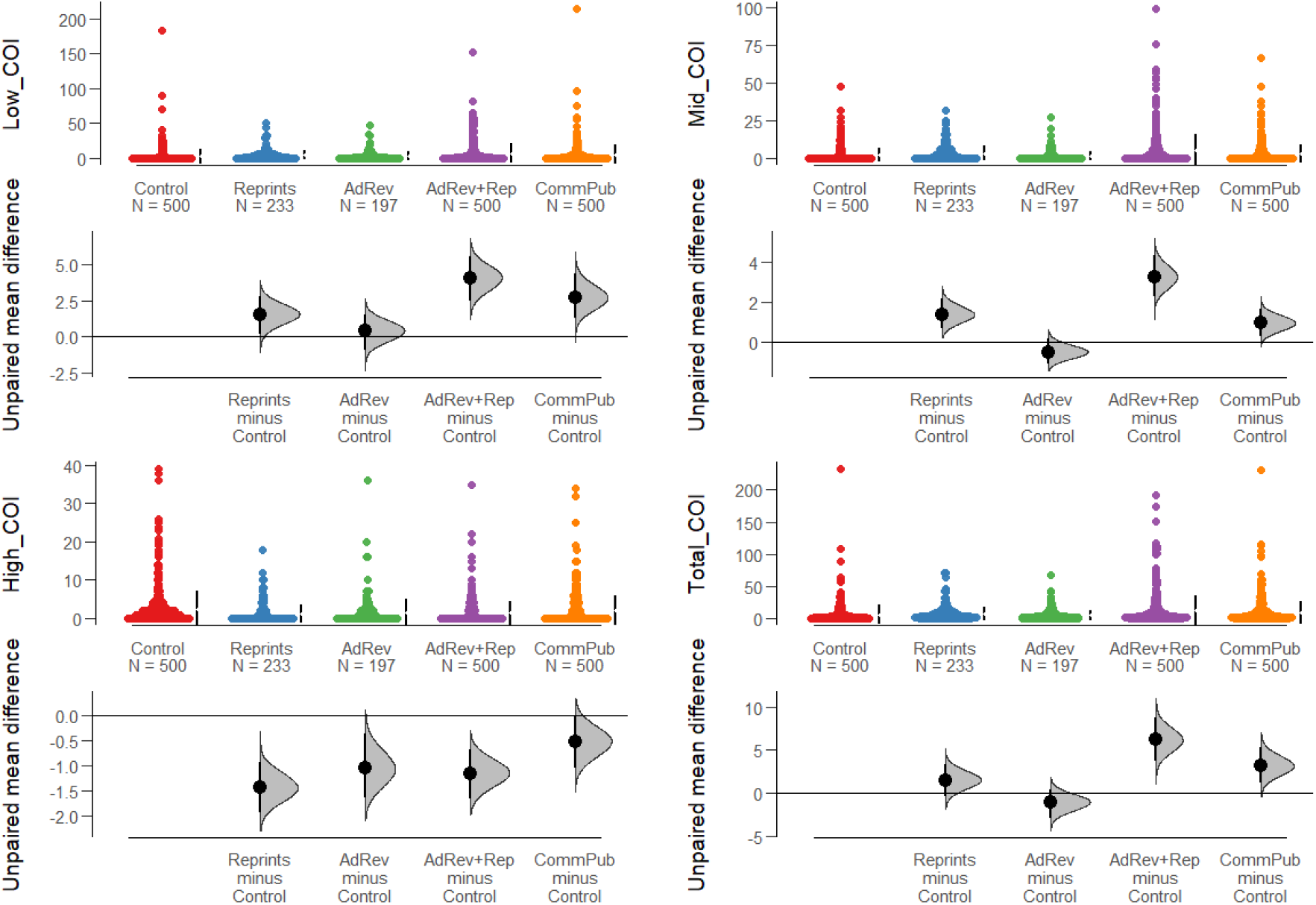
Shared-Control Estimation Plots for aCOI by level and total aCOI.

Nevertheless, certain patterns emerge when looking across tests. The data suggest there may be an aggregation effect whereby an increased number of commercial biases may result in a greater willingness to publish articles with higher aCOI rates. In each of the tests for total aCOI, low-level aCOI, and mid-level aCOI, journals that accept both ad revenue and reprints account for the greatest effects. A striking finding from this study is that in three of the four tests, the effect of advertising revenue, in isolation, is negative. In the remaining case, the effect size is negligible.

## Discussion

Editorial decision-making is a complex matter driven by a multitude of competing factors, many of which are not accounted for in the literature on the potential impacts of commercial biases. As biomedical publishing works to address these impacts within the context of broader concerns over COI and industry funding, it is imperative that new and revised policies are reflective of the best evidence available. The potential dangers of adopting new COI policies in the context of a dearth of evidence has become clear following the discovery of unanticipated and pernicious effects of COI disclosure.^32-33^ Disclosure statements have been shown to cause audiences to extend more trust to those holding conflicts of interest as disclosure provides an opportunity to display both honesty and expertise. Conflict disclosure can also lead to “moral licensing,” a phenomenon whereby those who disclose conflicts become unduly confident in their objectivity because transparency obligations have been fulfilled. In order to mitigate the risks of such unanticipated consequences in future policy proposals, recommendations must be based on a solid evidentiary foundation.

The results presented in this article work toward that end with respect to addressing commercial biases in biomedical publishing. One additional benefit of this study, compared to many others in the area, is that it includes non-clinical trial publications. As mentioned above, perspectives, comments, opinion pieces, and recruited articles are often selected solely on the basis of editorial discretion. As such, they may be especially open to commercial publishing biases. Additionally, the available data suggests that readers of biomedical journals are not always sensitive to the differences in peer- or editorially revised content. As such, these non-peer-reviewed publications may be exercising undue influence on practitioner understandings on the state of medical science.

Ultimately, the results presented here indicate that the presence of commercial biases predicts increases in industry favourability as measured by aCOI frequency and quantity. In particular, the data indicate that accepting reprint fees increases the likelihood that any given article published in a journal will have reported aCOI by a factor of 2.81. Additionally, these data show modest effects on the average quantity of aCOI in conflicted articles. That is, when journals accept both advertising revenue and reprint fees or belong to large commercial publishing firms, we see a modest increase in the average total aCOI per article. Interestingly, however, the results of the aCOI quantity analysis indicates that accepting advertising revenue, in isolation, has a modest negative effect on average aCOI per article. Finally, the data indicate that commercial publishing biases have a negligible, but negative, effect on average number of high-level aCOI per article.

Even though advertising revenue has been subject to the greatest scrutiny in the literature, it may represent the lowest cause for concern among the commercial biases evaluated in this study. This may indicate that something like the journalistic invisible wall is functioning appropriately in biomedical journals. Ultimately, these data indicate that the acceptance of fees for reprints may be the most impactful on commercial bias. In some respects, this makes sense. The potential for reprint revenue is the bias most directly tied to editorial decision-making. That is, the choice to publish a study favorable to industry, especially when that study might suggest new or expanded use of a drug, can be directly traced to reprint revenue.

Despite the suggestive nature of these findings, additional research should be conducted to verify and extend results. One important limitation of this study comes from low participation in Medline’s aCOI reporting program among many of the world’s top medical journals. Indeed, among the ten highest h-index medical journals, only one (*BMJ*) reports aCOI to Medline. Despite the above-mentioned limitations of disclosure statements, the availability of aCOI data has a real impact on our ability to evaluate potential risks. It would be helpful if more high-profile journals participated in Medline’s program. In the absence of such participation, supplementary research that collects data directly from target publications may be in order.

Additionally, further research should be conducted with respect to the impacts of commercial biases on other markers of editorial decision making. For example, researchers might take inspiration from the recent study identifying the prevalence of marketing trials across journals.^13^ Replicating this study with a data set stratified across journals representing a range of commercial biases would further add to the evidentiary foundation necessary to develop sound policies on commercial biases. New research might also use data on the prevalence of ghost authorship or improperly reported aCOI across journals to evaluate associations with commercial biases. In the meantime, the results presented here suggest that, as these data are being curated, attention should probably be focused on commercial publishing biases that can be tied most directly to editorial decision-making, specifically the collection of reprint revenues.

## Data Availability

Study data has been deposited with the Texas Data Repository

https://doi.org/10.18738/T8/VSWAJY

